# Factors influencing the motivation of maternal health workers in conflict setting of Mogadishu, Somalia

**DOI:** 10.1101/2022.01.10.22268888

**Authors:** Naima Said Sheikh, Abdi Gele

## Abstract

**Background:** Motivated health workers play an important role in delivering high-quality maternal health services, especially in low-income countries where maternal mortality rates are high, and shortages of human resource for health is prevalent. The aim of this study is to investigate maternal health workers’ motivation in three tertiary hospitals in Mogadishu Somalia.

**Method:** To investigate health worker motivation in Somalia, we used a semi-structured questionnaire that was validated and widely used in Sub-Saharan Africa. Data were collected from 220 health workers across three tertiary hospitals in Mogadishu between February and April 2020. Health worker motivation was measured using seven constructs: general motivation, burnout, job satisfaction, intrinsic job satisfaction, organizational commitment, conscientiousness, timeliness and attendance. A multivariate regression analysis was performed to determine the predictors of health worker motivation.

**Results:** The study found that male health workers have a higher work motivation, with a mean score of 92.75 (SD 21.31) versus 90.43 (SD 21.61) in women. A significant correlation was found between health workers’ motivation and being an assistant, nurse, physician, pediatric-assistant, midwife, supervisor and pharmacist. Unexpectedly, the gynecologists and midwives were the least motivated groups among the different professions, with mean scores of 83.63, (SD: 27.41) and 86.95 (SD: 21.08), respectively. Of the aforementioned seven motivation constructs, the highest mean motivation scores (from 1-5) were observed in conscientiousness and intrinsic job satisfaction.

**Conclusion:** The results highlight the importance of targeted interventions that increase female health workers’ motivation, particularly gynecologists and midwives in Somalia. This can be done by providing non-financial incentives, in addition to encouraging their participation in the decision-making process. Further research is needed to investigate the effect of a lack of motivation among gynecologists and midwives on maternal health in Somalia.

## Background

The large burden of mother and child mortality in Sub-Saharan Africa has been associated with inadequate health worker performance (1). According to the World Health Organization (WHO), African countries have the lowest health worker-to-population ratio in the world (2). An estimated shortage of 4.3 million health professionals was reported in low-income countries, and is more severe in Sub-Saharan Africa (3). Moreover, between 35 to 75% of available health workers are reported to be absent from clinics when on duty (4), while those who are present at work sometimes perform inadequately (1), failing to provide the correct diagnosis and treatment despite having the relevant medical knowledge (5).

In order to understand what motivates health workers, and how to improve their commitment and capacity to perform better, it is crucial to think of what health workers *know* and what they *do* in practice (6, 7). *Know* refers to the type of competencies and skills that enable people to do their job (8). Among other factors, this encompasses self-conception or professional identity, work orientation, self-confidence and self-regulatory skills, which all contribute to the accomplishment of the following goal adaptation (9). The *do* aspect of motivation refers to what extent health workers feel motivated and empowered to do their work tasks. This *do* is influenced by factors such as societal-, cultural-, personal values and personality tendencies, as well as factors related to organizational policies and management practices (9, 10).

Motivation is defined as “an individual’s degree of willingness to exert and maintain an effort towards organizational goals” (11). Motivation can be influenced by factors at different levels, such as the individual, organizational, health sectoral and community level (12-14). An individual’s motivation consists of two different types: intrinsic and extrinsic motivation. Intrinsic motivation refers to internal forces that motivate human agents, including health workers, to help meet their personal and organizational aims such as opportunities to use one’s ability, having self-expression, receiving appreciation and recognition. Extrinsic motivation includes substantive aspects such as working conditions, security, promotion and material benefits (12, 15). A health worker’s performance plays an important role in achieving a well-functioning health system, and also has an impact on the quality of the services provided (16). Hence, motivated health workers are considered to affect work performance, thereby increasing the quality, efficiency and equity of the healthcare services provided (11, 14).

Somalia has a population of 15.443 million people, with almost half (46.4%) under the age of 14 (17). The country experienced three decades of civil war beginning in 1991, which destroyed the healthcare system and health infrastructure in the country. Approximately 90% of current national spending priorities are spent on stability, security and administration, which means that there are few resources available for the healthcare sector and social services (20). The lack of an efficient public health system and the absence of proper regulation in the private sector, which is the dominant healthcare provider in Somalia, has made the situation worse, thus leading to a dire absence of regulatory checks in terms of efficiency and effectiveness (21). Inadequate health worker performance, an unregulated labor market and shortages of skilled health professionals have severely hampered further progress for Somalia’s healthcare system (19, 22).

Somalia’s maternal mortality rate (MMR) is 692 per 100,000, which is one of the highest in the world (23). The contributing factors for such a high MMR include socio-political instability and ongoing wars, as well as health system limitations (5). There were 27,000 women for every midwife and 30,000 people for every medical doctor in 2014 in Somalia (24). Traditional birth attendants (TBAs) are commonly used in delivery due to the shortage of skilled birth attendants (25). This means that many pregnant women do not receive skilled antenatal, delivery and postnatal care, which greatly increases their risk of dying from severe bleeding, infections or other preventable complications. For the last 10 years, a large number of medical doctors, nurses and midwives have graduated from local universities in Somalia. However, the potential of new graduates in reducing the human resource gap in the healthcare system, and subsequently improve the soaring MMR in the country, largely relies on health workers’ motivation and performance. The aim of this study is to investigate the motivation of maternal health workers, and the factors associated with their work performance, in Mogadishu, Somalia.

## Method and materials

A cross-sectional quantitative data was digitally collected between February and April 2020. Self-administered questionnaires (SAQs) were obtained from 220 health workers at the three largest maternal healthcare service providers in Somalia, namely the Benadir Maternity and Children’s Hospital, the Somali-Turkish Hospital and the SOS Hospital. The target population included assistants, nurses, midwives, pharmacists, physicians, pediatricians, gynecologists, and hospital supervisors working at maternal healthcare services. We ensured the diversity of the study participants by age, work position, education, gender, time spent at current profession and type of contract, including unpaid volunteers. To help fill the questionnaires, sufficient digital knowledge and access to digital technology were a prerequisite, as the SAQ was distributed digitally. Health workers who received the SAQ were encouraged to share it with their colleagues at the maternal health department.

We used an instrument developed by Mbindyo for the data collection (26). The instrument was validated with a good internal consistency, with a Cronbach’s Alpha of 0.75. Mbindyo’s tool was previously used in countries similar to Somalia such as Kenya (26) and Zambia (27). The instruments have 23 motivation items, and consisted of seven constructs: general motivation, burnout, job satisfaction, intrinsic job satisfaction, organizational commitment, conscientiousness and timeliness and attendance (26).

### Data analysis

Data was entered into Microsoft software, and then exported to statistical software STATA, version 16 for analysis. A Likert scale of one to five was entered, in which a score of “5” represented strongly disagree for positively worded statements. The negatively worded statements were coded “1”, so that a score of “5” indicated strongly agree as marked in bold in Table 2. The mean for the negative statements was then recalculated with a reverse coding; therefore, a higher mean for all items indicates a higher motivational outcome. The total of 23-item motivation scores was used to provide summary statistics. Afterwards, we adopted the simplified 10-item index suggested by Mbindyo (26), which is easier and more suitable for measuring healthcare motivation in Sub-Saharan Africa.

A multiple linear regression analysis was conducted to estimate the relationship between the dependent variables and independent variables, and to identify whether motivation scores differ between gender, age group, hospital, the type of contract and time spent in current workplace.

Since organizational commitment was one of the dependent variables, hospitals were included in the analysis as an independent variable. Health workers at the three hospitals had different employment contracts, including volunteers, contract staff (working for NGOs) and government employee. All three contract types were included in the analysis. This allowed us to determine whether motivation differed across hospital and employment status.

The outcome variable was the total motivation score, which was calculated by the sum of the scores of the 23 items in the motivation scales. Finally, the total of the simplified 10-item motivation scores was entered into the analysis. A p-value of ≤0.05 was considered significant.

### Ethical issues

The study protocol followed the principles outlined in the Declaration of Helsinki. The confidentiality of the participants’ responses was guaranteed to everyone, with verbal consent obtained from participants after being informed about the study objectives. The data were stored, analyzed and reported in a way that does not allow for the identification of the health workers who participated in the study.

## Results

Table 1 shows the demographic characteristics of health workers across the three hospitals. The vast majority of the study participants were between the ages of 20-29 years (75%), while only 5% were >40 years. There was no significant age difference between men and women, and the majority had a Bachelor’s degree (90%). Men were more likely to have a Master’s degree (14%) than women (6%). Regarding contract type, more men had a government contract (39%), thereby giving them more economic and job security. Women were more likely to work for NGOs (40%). Most of the participants worked at their current position for ≤ 5 years (74%).

**Table 1:** General characteristics of study participants by gender.

|  | Variables | Male (N=77) | Female (N=143) | Total (N=220) |
| --- | --- | --- | --- | --- |
|  |  | N (%) | N (%) | N (%) |
| Age | 20-29 | 59 (77) | 105 (73) | 164 (75) |
|  | 30-39 | 15 (19) | 30 (21) | 45 (20) |
|  | 40-49 | 1 (1) | 4 (3) | 5 (2) |
|  | >50 | 2 (3) | 4 (3) | 6 (3) |
| Education level | High school | 0 | 2 (1) | 2 (1) |
|  | Bachelor's degree | 66 (86) | 132 (92) | 198 (90) |
|  | Master's degree | 11 (14) | 9 (6) | 20 (9) |
| Type of contract | Government employment | 30 (39) | 48 (34) | 78 (35) |
|  | Contract staff | 26 (34) | 57 (40) | 83 (38) |
|  | Voluntary staff | 21 (27) | 38 (27) | 59 (27) |
| Place of work | Benadir Hospital | 51 (66) | 66 (46) | 117 (53) |
|  | Somali-Turkish Hospital | 16 (21) | 43 (30) | 59 (27) |
|  | SOS Hospital | 10 (13) | 34 (24) | 44 (20) |
| Years worked in current profession | < a year | 21 (27) | 46 (32) | 67 (30) |
|  | 1–5 years | 36 (47) | 61 (43) | 97 (44) |
|  | 6–10 years | 11 (14) | 19 (13) | 30 (14) |
|  | > 10 years | 9 (12) | 17 (12) | 26 (12) |
| Profession | Assistant | 5 (6) | 2 (1) | 7 (3) |
|  | Nurse | 18 (23) | 47 (33) | 65 (30) |
|  | Physician | 23 (30) | 28 (20) | 51 (23) |
|  | Midwife | 0 | 37 (26) | 37 (17) |
|  | Pharmacist | 2 (3) | 4 (3) | 6 (3) |
|  | Pediatric assistant | 19 (25) | 1 (39) | 32 (15) |
|  | Pediatrician | 6 (8) | 1 (1) | 7 (3) |
|  | Gynecologist | 1 (1) | 7 (5) | 8 (4) |
|  | Supervisor | 3 (4) | 4 (3) | 7 (3) |

### Motivational status

Regarding intrinsic job satisfaction, and using a scale from 1 to 5 (1=fully demotivated and 5=fully motivated), we found a mean score of 4.70 (SD: 0.54) and 4.68 (SD: 0.59) in items 9 and 10, respectively, thereby indicating that health workers are highly satisfied with their job opportunities and accomplishments (Table 2).

**Table 2:** Summary statistics for motivation outcome constructs.

| Construct | No. | Statement | Mean | SD |
| --- | --- | --- | --- | --- |
| General motivation | 1 | These days, I feel motivated to work as hard as I can* | 4.33 | 0.95 |
|  | 2 | **I only do this job so that I get paid at the end of the month** | 4.01 | 1.27 |
|  | 3 | **I do this job, as it provides long-term security for me** | 2.10 | 1.04 |
| Burnout | 4 | **I feel emotionally drained at the end of every day** | 2.55 | 1.30 |
|  | 5 | **Sometimes when I get up in the morning, I dread having to face another day** | 1.81 | 1.00 |
| Job satisfaction | 6 | Overall, I am very satisfied with my job* | 4.65 | 0.64 |
|  | 7 | **I am not satisfied with my colleagues in my ward** | 3.85 | 1.29 |
|  | 8 | I am satisfied with my supervisor | 4.29 | 0.88 |
| Intrinsic job satisfaction | 9 | I am satisfied with the opportunity to use my abilities in my job* | 4.68 | 0.54 |
|  | 10 | I am satisfied that I accomplish something worthwhile in this job* | 4.70 | 0.59 |
|  | 11 | **I do not think that my work in the hospital is valuable these days** | 4.24 | 0.98 |
| Organizational commitment | 12 | I am proud to be working for this hospital* | 4.59 | 0.74 |
|  | 13 | I find that my values and this hospital's values are very similar | 3.79 | 1.21 |
|  | 14 | I am glad that I work for this facility, rather than other facilities in the country* | 2.91 | 1.18 |
|  | 15 | **I feel very little commitment to this hospital** | 3.18 | 1.19 |
|  | 16 | This hospital really inspires me to do my very best on the job* | 4.44 | 0.99 |
| Conscientiousness | 17 | **I cannot be relied on by my colleagues at work** | 4.25 | 0.99 |
|  | 18 | I always complete my tasks efficiently and correctly* | 4.65 | 0.60 |
|  | 19 | I am a hard worker* | 4.61 | 0.65 |
|  | 20 | I do things that need doing without being asked or told | 4.67 | 0.62 |
| Timeliness and attendance | 21 | I am punctual about coming to work* | 4.57 | 0.66 |
|  | 22 | **I am often absent from work** | 4.39 | 0.98 |
|  | 23 | **It is not a problem if I sometimes come late to work** | 4.00 | 1.12 |
| SD: Standard Deviation. Mean scores (1-5) 1 (Strongly disagree) to 5 (Strongly agree). The simplified 10 items are marked with an asterisk (*). Negatively worded statements are in Bold. The scale for negatively worded statements was 1 (strongly agree) to 5 (strongly disagree). Thus, a high score shows a disagreement with a negative statement, and therefore indicates a higher motivation. |  |  |  |  |

The results in Tables 3 and 4 show that male health workers have slightly higher overall mean motivation scores (92.75, SD: 21.31 for men) than female health workers (mean score: 90.43, SD: 21.61). In terms of profession, assistants were the most motivated group, with a mean score of 95.85 (SD:16.43), followed by physicians (Mean: 94.41, SD: 19.48), while pharmacists and supervisors had a lower mean score of 92.23 (SD: 22.47). Unexpectedly, gynecologists and midwives were the least motivated groups (mean score: 83.63, SD: 27.41 and 86.95, SD: 21.08, respectively).

**Table 3:**
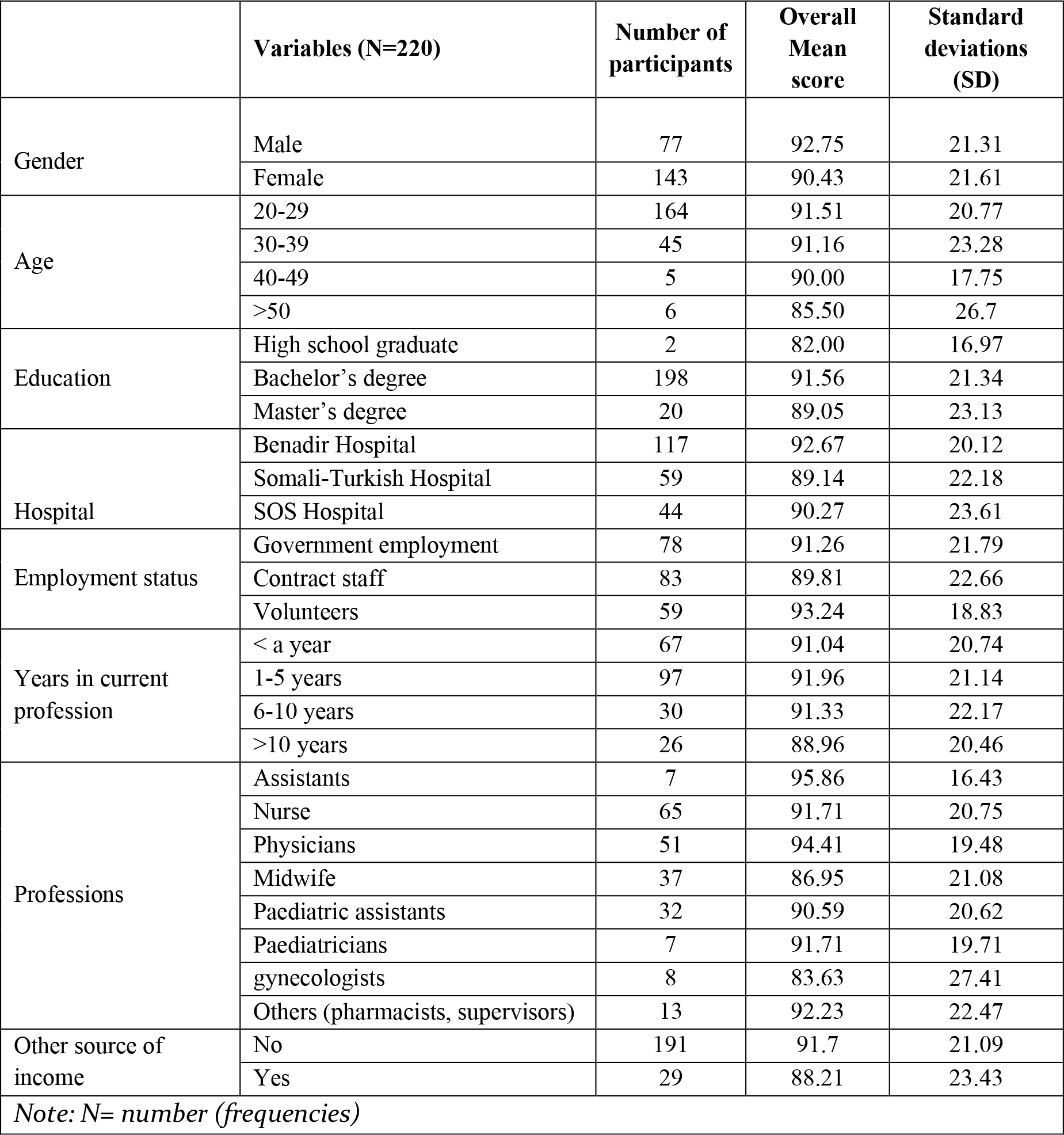
Overall motivation scores of health professionals by socio-demographic characteristics.

**Table 4:** Mean motivation scores (1-5) by hospital and gender.

| Motivation items | Benadir Maternity Hospital<br>(N = 117) |  | Somali-Turkish Hospital<br>(N = 59) |  | SOS Hospital<br>(N = 44) |  |
| --- | --- | --- | --- | --- | --- | --- |
|  | Male<br>(N = 51) | Female<br>(N = 66) | Male<br>(N = 16) | Female<br>(N = 43) | Male<br>(N = 10) | Female<br>(N = 34) |
| General motivation | 11.18 | 10.55 | 10.13 | 9.47 | 9.8 | 10.71 |
| Burnout | 4.67 | 4.17 | 4.69 | 4.26 | 3.8 | 4.5 |
| Job satisfaction | 13.47 | 12.94 | 12.19 | 12.16 | 12.8 | 12.53 |
| Intrinsic job satisfaction | 13.86 | 13.73 | 14.00 | 13.37 | 13.4 | 13.21 |
| Organizational commitment | 19.94 | 18.85 | 18.31 | 17.84 | 19.4 | 18.91 |
| Conscientiousness | 17.98 | 18.38 | 17.88 | 18.37 | 18.8 | 17.74 |
| Attendance and timeliness | 12.8 | 13.11 | 13.06 | 13.26 | 12.9 | 12.5 |
| **Overall mean scores** | **93.90** | **91.73** | **90.26** | **88.73** | **90.90** | **90.09** |

Regarding type of contract, government employees (Benadir Maternity Hospital) had a higher motivation compared to those with contract staff. Volunteers, government employees and contract staff had mean scores of 93.24 (SD: 18.83), 91.26 (SD: 21.79) and 89.81 (SD: 22.66), respectively.

As shown in Table 4, male health workers had a slightly higher general motivation, better job satisfaction, better organizational commitment and intrinsic job satisfaction in all hospitals, with the exception SOS, where male participants had a lower general motivation and burnout. In the SOS Hospital, females had higher scores on general motivation and burnout. Even though men were more motivated, female health workers at the Benadir and Somali-Turkish Hospitals had a higher motivation in relation to conscientiousness, attendance and timeliness.

The regression analysis in Table 5 shows variations in motivation between health workers across education, profession, hospital, type of employment, age and time spent at current profession.

**Table 5:**
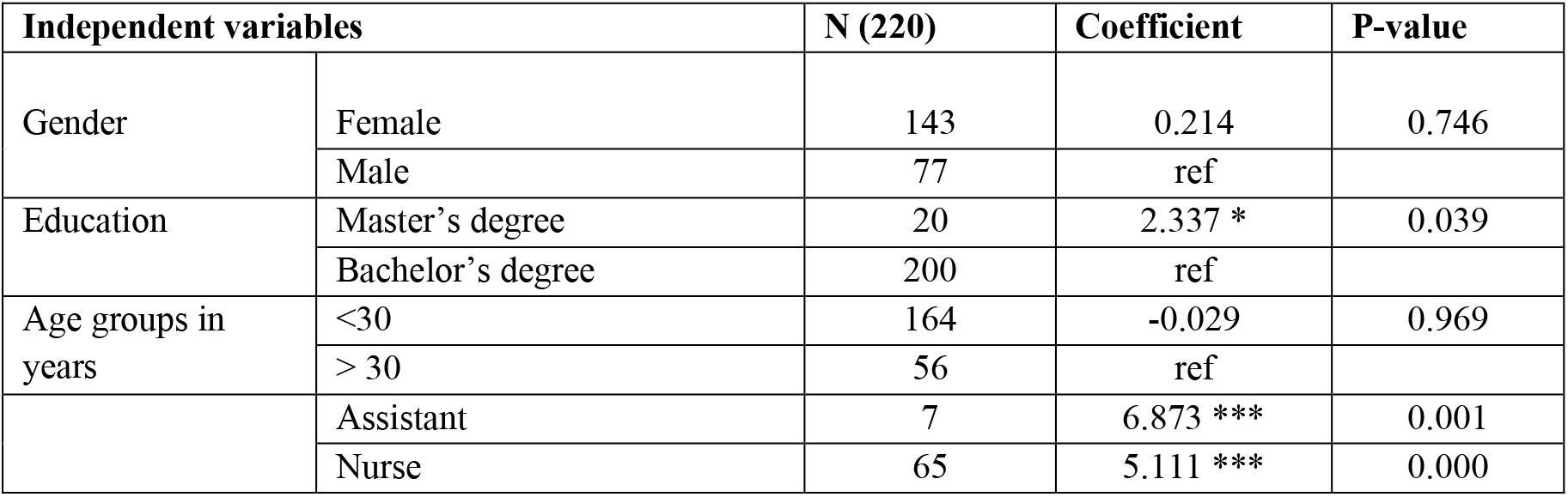
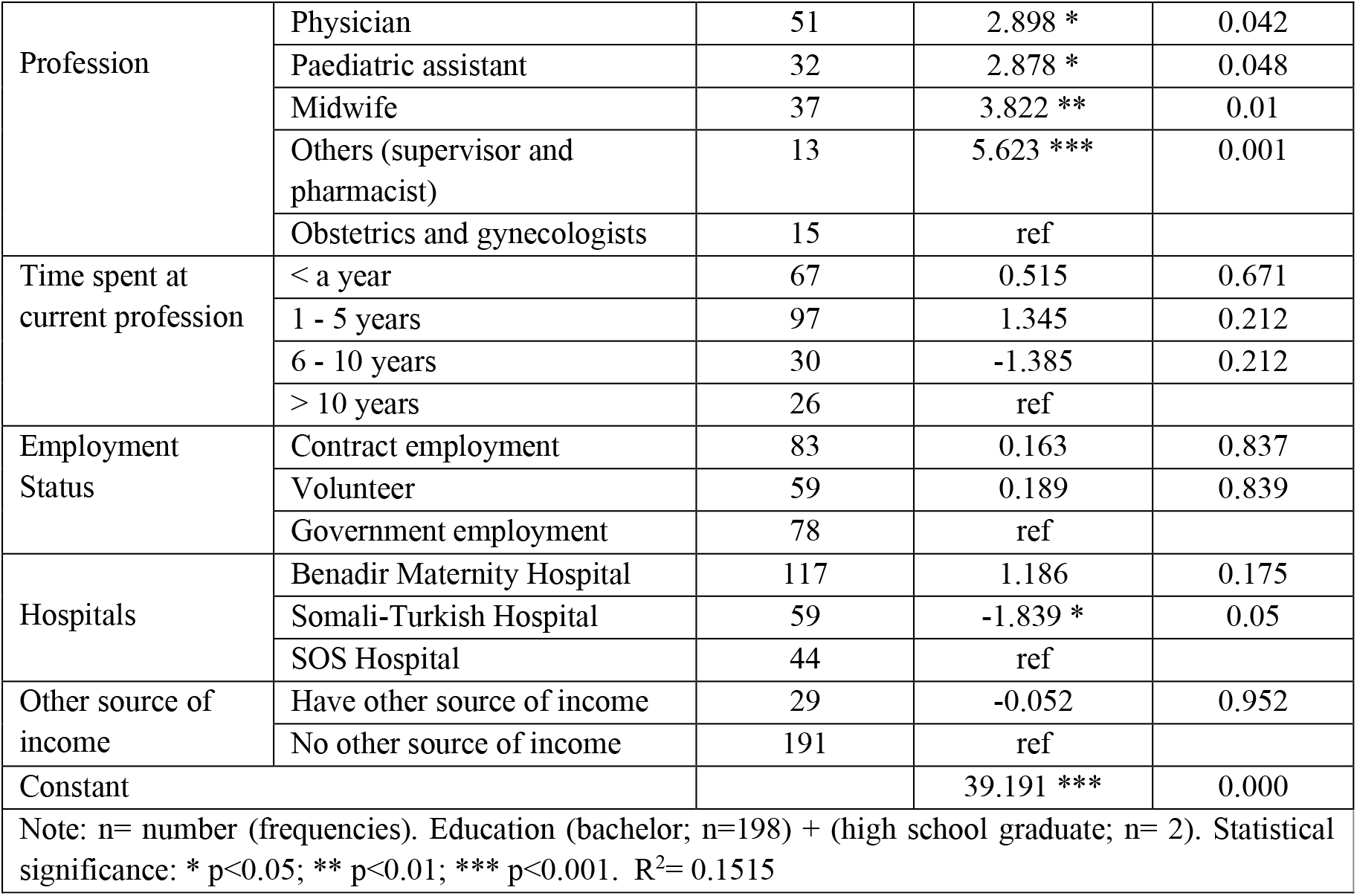
Linear Regression Model for Independent Variables and the 10-Item Motivation Scores.

Regarding education and profession, a higher education (Masters’ degree) increases the level of motivation compared to those with a lower level of education (bachelor and high school). Professions such as assistants, nurses, physicians, pediatric assistants, midwives and others (supervisors and pharmacists) had a significantly higher motivation compared to obstetricians and gynecologists. Regarding hospitals, health workers at the Somali-Turkish Hospital seem to be significantly less motivated than health workers at the SOS Hospital (b: -1.839, p-value: 0.05).

## Discussion

This study investigates health workers’ motivation in conflict-affected settings in Somalia. Motivated health workers are seen to be important for the provision of quality health services and improved work performance. Our findings show that Somali health workers are more motivated than the health workers in Kenya (30), Tanzania (11), Zambia (31) and Mali (32). The reason for this could be because Somali health workers live in a conflict setting where finding a job is extremely difficult; their motivation may therefore be influenced by their perception that “they are so blessed” to have a job; a situation called a “frame of reference effect.”

Overall, this study revealed that male health workers are more motivated than female health workers, whereas studies in Ethiopia and Zambia found that females were more motivated than males (31). The difference between our study and other studies could be explained by a patriarchal culture in Somalia, where females experience a significant cultural oppression in employment across all sectors, including the healthcare sector. As seen in Table 3, male health workers had higher positions in the hospital across several professions, i.e., physicians, pediatric assistants and pediatricians (specialists). More male health workers had a Master’s degree (specialist education) than females, which was significantly correlated with staff motivation. This explains why males are more motivated than females. Furthermore, more males had government employment, while contract staff (temporary NGO staff) was more common among females. The contract staff normally experienced job insecurity, as the NGOs they work for may leave the country overnight. In addition, NGOs’ staff do not have a regular salary, while government employees have sustainable jobs with a higher and reliable salary, as well as other opportunities that provide them with career development, such as scholarships.

This study reveals that there is a significant relationship between health workers’ motivation and their profession. In agreement with our findings, prior studies found that the availability of training and educational opportunities for health workers, as well as improving their knowledge and skills, influenced their motivation and work performance (11). Our study shows that gynecologists are less motivated compared to specialties such as midwives. Gynecologists are critical in maternal health in Somalia, where such specialties are scarce and maternal mortality is high. The impact from multitasking, a high intensity workspace and high expectations from leadership and patients might have led to some degree of physical or emotional exhaustion, and subsequent lack of motivation for gynecologists. However, the reasons behind the lack of motivation of gynecologists in Somalia require further exploration.

Surprisingly, volunteers have the highest motivation score compared to salaried employees. This implies that the salary is not the primary motivating factor for employees, but rather that inner motivation influences health workers’ motivation. This suggests that constructs concerning inner motivation are of more importance than constructs concerning other aspects of motivation. Previous studies reported that the individual determinant of motivation among East African health workers resulted from the intrinsic desire to help others (family, community or the public) (33). The reason why more volunteers were working at the hospitals studied could be the prevalent unemployment in the country, and the fact that unemployed health professionals consider voluntary work to be a better option than staying home and not practicing their academic career. Volunteerism also provides an experience that is a prerequisite for the possibility of future employment at healthcare facilities.

One of the limitations of this study is the safety issue in Mogadishu, where health workers might not freely express their feelings, which could result in biased information. Another limitation is that it was not possible to verify the working conditions in Somalia and the availability of essential medical equipment and staff during the data collection. A fieldwork, including observation research, would have provided a more insightful analysis. This study was conducted in three healthcare institutions in Mogadishu; hence, the staff motivation in other healthcare facilities in the country may be different. However, the three hospitals in our study are the biggest tertiary hospitals in Somalia, and they serve not only the inhabitants of Mogadishu, but also patients from all corners of Somalia. Although we did not use a random sampling technique, the results from this study can be extrapolated to the general healthcare system in Somalia, since many healthcare workers in other hospitals and clinics may share similar characteristics with our study participants.

## Conclusion

The findings of this study are crucial for improving the health workers’ motivation and performance in Somalia, which is essential in achieving UN Sustainable Development Goals (especially Target 3.1). Improving recruitment strategies, the provision of flexible employment opportunities, improving gender-based equalities, fostering teamwork and encouraging staff to innovate are critical in the motivation of healthcare staff in Somalia. These suggestions were also mentioned in the World Health Report on preparing the workforce (34). Our study focuses on provider behaviors, so we therefore recommend future research that investigates the demand side perspective. Studying from both supply and demand could provide Somalia with better strategies when applying interventions in the healthcare system in general, and improving the quality of healthcare provided.

## Data Availability

All data produced in the present study are available upon reasonable request to the authors

## Acknowledgments

We would like to thank the study participants for their time and participation in the study.

## References

1. Alexander K Rowe, Don de Savigny, Claudio F Lanata, Cesar G Victora. How can we achieve and maintain high-quality performance of health workers in low resource settings?, The Lancet, Volume 366, Issue 9490, 2005, Pages 1026–1035.

2. Kofi Aduo-Adjei, Emmanuel Dadzie Odoom, Opoku Mensah Forster. The Impact of Motivation on the Work Performance of Health Workers (Korle Bu Teaching Hospital): Evidence from Ghana. 2016. 1(2):45–50.

3. World Health Organization 2006: Working together for health. The World Health Report Geneva, 2006.

4. Chaudhury, N., Hammer, J., Kremer, M., Muralidharan, K., & Rogers, F. H. (2006). Missing in action: Teacher and health worker absence in developing countries. The Journal of Economic Perspectives: A journal of the American Economic Association, 20(1), 91–116. https://doi.org/10.1257/089533006776526058.

5. Das, Jishnu, Jeffrey Hammer, and Kenneth Leonard. 2008. “The Quality of Medical Advice in Low-Income Countries.” Journal of Economic Perspectives, 22 (2): 93–114. DOI: 10.1257/jep.22.2.93.

6. Mohanan M, Vera-Hernandez M, Das V, et al. The know-do gap in quality of health care for childhood diarrhea and pneumonia in rural India. JAMA Pediatr 2015;169:349–57.

7. Leonard KL, Masatu MC. Professionalism and the Know-do Gap: Exploring Intrinsic Motivation among Health Workers in Tanzania. Health Econ 2010;19:1461–77.

8. Fritzen SA. Strategic management of the health workforce in developing countries: What have we learned? Hum Resour Health. 2007 Feb 26;5:4. doi: 10.1186/1478-4491-5-4.

9. Hotchkiss DR, Banteyerga H, Tharaney M. Job satisfaction and motivation among public sector health workers: Evidence from Ethiopia. Hum Resour Health. 2015 Oct 29;13:83. doi: 10.1186/s12960-015-0083-6.

10. Franco LM, Bennett S, Kanfer R, Stubblebine P. Determinants and consequences of health worker motivation in hospitals in Jordan and Georgia. Soc Sci Med. 2004 Jan;58(2):343–55. doi: 10.1016/s0277-9536(03)00203-x.

11. Dagne, T. Beyene, W. & Berhanu, N. Motivation and Factors Affecting It among Health Professionals in the Public Hospitals, Central Ethiopia. 2015.

12. Leonard, Kenneth L.; Masatu, Melkiory C.; Herbst, Christopher H.; Lemiere, Christophe. 2015. The Systematic Assessment of Health Worker Performance : A Framework for Analysis and its Application in Tanzania. Health, Nutrition and Population Discussion Paper; World Bank.

13. Weldegebriel Z, Ejigu Y, Weldegebreal F, Woldie M. Motivation of health workers and associated factors in public hospitals of West Amhara, Northwest Ethiopia. Patient Prefer Adherence. 2016 Feb 15;10:159–69.

14. Khim K. Are health workers motivated by income? Job motivation of Cambodian primary health workers implementing performance-based financing. Glob Health Action. 2016;9:31068. Published 2016 Jun 17.

15. Baljoon, R. Banjar, H. & Banakhar, M. Nurses’ Work Motivation and the Factors Affecting It: A Scoping Review. 2018. International Journal of Nursing & Clinical Practices. 5: 277.

16. Bradley, S., McAuliffe, E. Mid-level providers in emergency obstetric and newborn health care: Factors affecting their performance and retention within the Malawian health system. Hum Resour Health 7, 14 (2009). https://doi.org/10.1186/1478-4491-7-14.

17. United Nations, 2019, United Nations data Somalia.http://data.un.org/en/iso/so.html.

18. World Health Organization. Somalia: Building a stronger primary health care system. 2020. https://www.who.int/news-room/feature-stories/detail/somalia-building-a-stronger-primary-health-care-system.

19. Anderson, J L. 2009, THE MOST FAILED STATE Is Somalia’s new President a viable ally? The New Yorker. https://www.newyorker.com/magazine/2009/12/14/the-most-failed-state.

20. World Bank. The World Bank In Somalia, Overview, 2019. https://www.worldbank.org/en/country/somalia/overview.

21. Regional Health System Observatory 2006, HEALTH SYSTEM PROFILE, SOMALIA Regional Health Systems Observatory. WHO, EMRO.

22. Mitesh, M. Understanding Somalia: Reasons for its State Failure. 2015. http://www.diplomatmagazine.nl/2015/01/04/understanding-somalia-reasons-state-failure%EF%BB%BF%EF%BB%BF%EF%BB%BF/.

23. The Federal Republic of Somalia. The Somali Health and Demographic Survey 2020.

24. The WHO Regional Office for the Eastern Mediterranean. Somalia crisis: Health update. WHO 2015. Available; http://www.emro.who.int/images/stories/eha/documents/infographic-somalia-health_sector_update.pdf?ua=1.

25. Orya, E., Adaji, S., Pyone, T. et al. Strengthening close to community provision of maternal health services in fragile settings: An exploration of the changing roles of TBAs in Sierra Leone and Somaliland. BMC Health Serv Res 17, 460 (2017). https://doi.org/10.1186/s12913-017-2400-3.

26. Mbindyo PM, Blaauw D, Gilson L, English M. Developing a tool to measure health worker motivation in district hospitals in Kenya. Hum Resour Health. 2009 May 20;7:40.

27. Mutale W, Ayles H, Bond V, Mwanamwenge MT, Balabanova D. Measuring health workers’ motivation in rural health facilities: Baseline results from three study districts in Zambia. Hum Resour Health. 2013 Feb 21;11:8.

28. Dieleman, M. Toonen, J. Toure, H. & Martineau, T. The match between motivation and performance management of health sector workers in Mali. 2006. Human Resources for Health 4:2 Pdf. P. 3.

29. Muthuri RNDK, Senkubuge F, Hongoro C. Determinants of Motivation among Healthcare Workers in the East African Community between 2009-2019: A Systematic Review. Healthcare (Basel). 2020;8(2):164. Published 2020 June 10. doi:10.3390/healthcare8020164.

30. World Health Organization. Working together for health. The World Health Report Geneva. 2006. https://www.who.int/workforcealliance/knowledge/resources/whreport_2006/en/.

